# Cutaneous leishmaniasis in British troops following jungle training in Belize: cumulative incidence and potential risk practices

**DOI:** 10.1101/2024.01.30.24302002

**Authors:** Ngwa Niba Rawlings, Mark Bailey, Peter Craig, Orin Courtenay

## Abstract

**Introduction:** British soldiers undergoing jungle training (JT) in Belize typically experience a relatively low risk of developing cutaneous leishmaniasis (CL). However, an uncharacteristically large outbreak of CL occurred in 2022. This study aimed to determine the cumulative incidence (CUMI) of the disease and highlight potential shortcomings in personal protective measures (PPMs) to mitigate exposure to sand fly vector bites.

**Methods:** A retrospective analysis was conducted on medical records of CL cases between 2005 and 2022, as well as on questionnaire responses regarding PPMs administered to CL cases in 2022. Data were sourced from Defence Public Health Unit, Military Environmental Health Department and British Army Training Support Unit Belize.

**Results:** Eighty-one confirmed clinical CL cases were recorded between 2005 and 2022, with a substantial peak (38 cases) in 2022. Most cases occurred during the wet season. Pre-2022, the median CUMI per 8-week deployment was 0.90% (Q1–Q3: 0.34%–1.34%), with an annual variation of 0.2% to 2.0%. In 2022, the CUMI spiked to 4.22%, associated with a risk ratio of 5.3 (95% C.I.s: 3.41, 8.16), and rising to a CUMI of 7.3% in a single unit of 450 men (33 CL cases) in late 2022. These values are significantly higher than the median CUMI of all previous years, and to published reports for other CL-endemic regions. Troop responses identified limitations in the supply of optimal equipment, and in sand fly bite and leishmaniasis risk avoidance information provided by the pre-deployment health education (PDHE) programme. Compliance with PDHE advise was also suboptimal, with irregular use of insect repellents, protective clothing / head netting, and insecticide-treated hammocks.

**Conclusions:** The reasons behind the unusually high numbers of CL cases and CUMI in 2022 remain unclear, emphasising the need to improve PPM provision and implement a comprehensive PDHE programme for troops undergoing JT in Belize.

## 1. Introduction

Cutaneous leishmaniasis (CL) is the most common form of human leishmaniasis. *Leishmania* are protozoan parasites transmitted by infectious female Phlebotomine sand flies when blood-feeding on human hosts [1,2]. Clinical signs of CL predominantly presents as skin lesions, particularly ulcers, and can significantly impact an individual’s quality of life [3,4]. CL is prevalent in the Americas, Mediterranean basin countries, sub-Saharan Africa, the Middle East, and central and south east Asia, amounting to an estimated annual incidence of 600,000 to 1 million cases worldwide [2,5]. In the Americas, CL is endemic in 19 countries where 1,105,545 cases were reported between 2001 and 2021, with an average of 52,645 cases per year [6].

CL is commonly observed among soldiers who have returned from international deployments to CL endemic regions. Notable case numbers are associated with deployments to Iraq [7,8], Afghanistan [7–9], French Guiana [10,11], Brazil [12,13], Panama [14], Peru [15], and Belize [16–19], with attack rates ranging from 2.1% to 25.2%. The risk of sand fly bite exposure leading to infection has been attributed to several factors, including failure to adopt personal protective measures (PPMs) [10,14,15,18,20–25], participation in military training activities during periods of sand fly biting [10,14,15], and in marshy areas near streams or rivers [20,23], and operations in areas undergoing significant environmental changes including deforestation, illegal logging, and other land use alterations for agriculture [10,15].

Leishmaniasis usually impedes soldiers’ ability to perform their duties, leading to increased sick leave, reduced combat readiness, and potential long-term health consequences. Furthermore, the need for diagnosis, treatment, and compensation claims further strain military resources. Between 1998 and 2021, >65% of diagnosed CL cases in the military returning to the UK were recently deployed to Belize (formerly British Honduras) [16], where 306 CL cases were recorded in British soldiers between 1978 and 1990 [17]. During a more recent outbreak in Belize in 2022, 33 CL cases were confirmed in British soldiers participating in a single 8-week jungle exercise (MOD data – restricted access). In addition to military jungle training (JT), infections leading to CL in Belize are predominantly associated with forest activites including chewing-gum latex collection, and tourism [16,26–28].

Despite the serious impacts of CL, there is a notable gap in the current knowledge amongst military personnel concerning the epidemiology, incidence, and risk factors for exposure to potentially infectious sand fly vectors. To address these knowledge gaps, this study undertook a review of the cumulative incidence of CL cases arising from British military JT in Belize between 2005 and 2022, and conducted a preliminary investigation of compliance to provide and adopt personal protective measures (PPMs) to mitigate vector exposure during deployment.

## 2. Methods

### Ethics statement

The study was conducted in accordance with the principles of the Declaration of Helsinki and all applicable laws and regulations governing research ethics. The study was a review of anonymised secondary data belonging to, and authorised, by the Ministry of Defence.

### Study setting

British troops undergo JT centred around the British military base in the British Army Training Support Unit Belize (BATSUB). The training areas include Rio Bravo, Laguna Sector, Gallon Jug and Yalbac in the north west of the country; 1963 Line, Chiquibul (Guacamallo) and Mountain Pine Ridge in the south west; Baldy Beacon and Sibuan Gorge in the centre, and Manatee north in the east (Fig 1).

**Figure 1:**
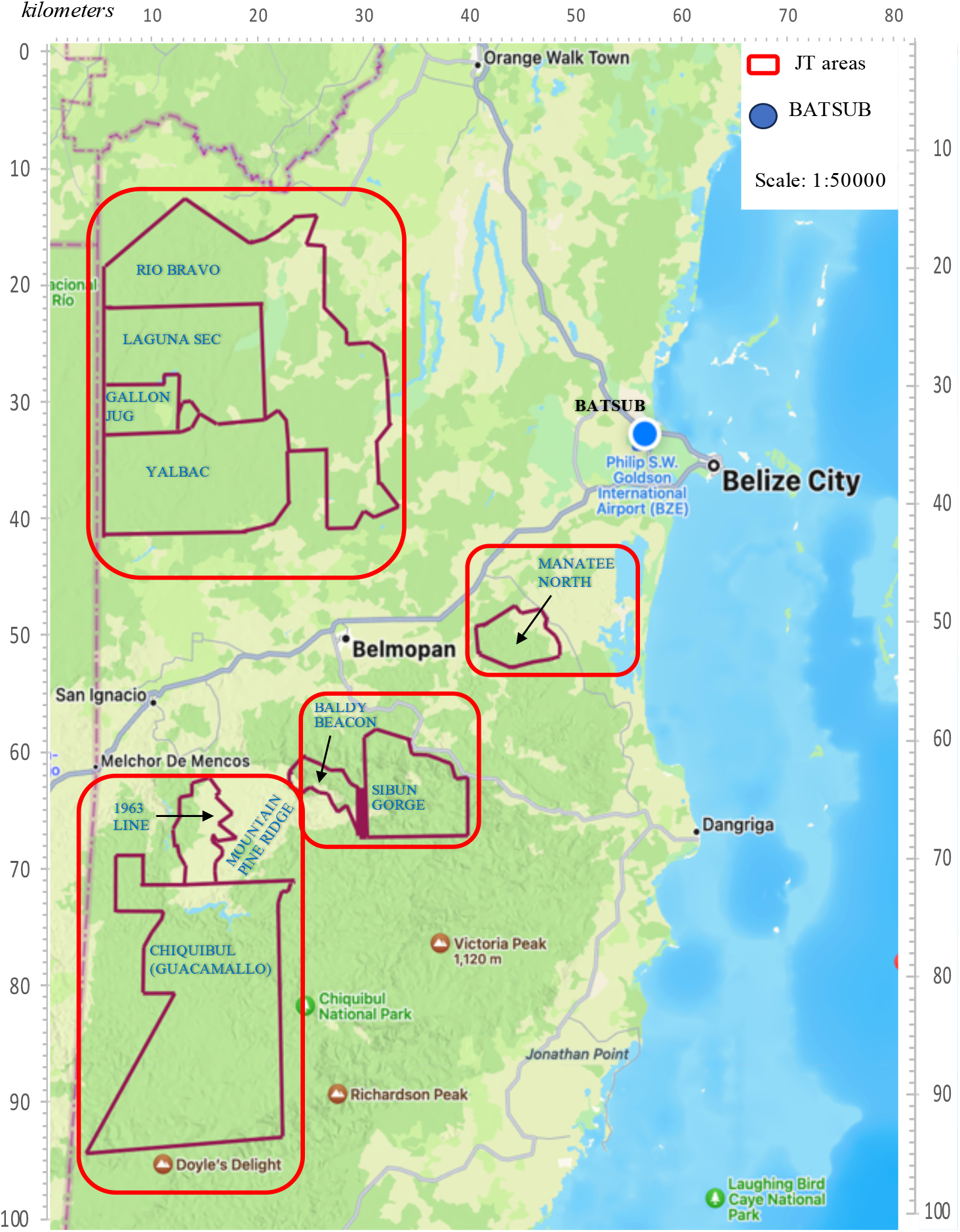
British army training areas (red boxes) and BATSUB in Belize, Central America.

These training grounds are characterised by tropical rainforests, predominantly consisting of primary and/or secondary jungle including hardwoods, palms, pines and various flowering plants. Belize experiences two distinct climatic seasons: a dry season from December to April and a wet season from May to November, with average temperatures ranging from 23°C to 27°C throughout the year. In the wet season, the southern part of the country receives an average monthly rainfall of 150 mm to 400 mm, while other areas receive less than 100 mm per month [29,30]. Troops are deployed in all seasons.

CL is endemic in Belize [31], primarily due to infection with *Leishmania (L) mexicana* and *L*.*(V*.*) braziliensis* [16,32,33]. Multiple species of forest-dwelling sand fly vectors are historically documented [34,35] and rodents are suspected to be the zoonotic reservoir hosts [36–39]. Current understanding of *Leishmania* transmission in Belize relies on historical accounts that documented CL prevalence in the local human population [40], the diversity and abundance of rodent [35,39–41] and sand fly [35,36] species, their associated phenologies, ecotypes and sand fly biting behaviours [35,36], and attempts to detect *Leishmania* infections to incriminate these species as reservoirs and vectors, respectively.

### Study population

The British military’s JT program in Belize involves approximately 1000 soldiers each year comprising various UK units, being sent to BATSUB, where they undergo an 8-week program designed to train soldiers in the jungle environment. The programme comprises two weeks of acclimatisation and preparation in BATSUB, five weeks of uninterupted time living in the jungle undergoing training (jungle Ex phase) based at one of the indicated training areas (Fig 1), followed by a week of rest and recuperation (R&R) housed back at BATSUB. Participating in these training exercises are ranks of junior non-commissioned officers (JNCO; Privates, Lance Corporals, and Corporals), senior non-commissioned officers (SNCO; Sergeants, Staff Sergeants, and Warrant Officers), and Officers. Soldiers should receive pre-deployment health education (PDHE) providing them with information regarding health risks in the deployment area, highlighting the importance of adopting and usage of PPMs and related hygiene practices. The goal is to equip soldiers with the knowledge and skills needed to maintain their well-being. Typically, PDHE is intended to be delivered by qualified military environmental health practitioners (EHPs); however, there are instances when it is delivered by combat medical technicians (CMTs).

### Data collection and Analyses

Anonymised medical records of clinically and laboratory confirmed CL cases among British soldiers who underwent JT in Belize between 2005 and 2022 were provided by the Defence Public Health Unit; the total numbers of British soldiers deployed (at risk) during specified periods of JT were provided by BATSUB. No duplicate/missing records were identified. The cumulative incidence (CUMI) of CL was calculated by dividing the number of new CL cases during specified periods of training by the total number of troops deployed (all condiered at risk) at the beginning of the 8-week training period (Tables 2 & 3). The annual CUMI was calculated by aggregating annual total CL case and deployed troop numbers (Table 2).

**Table 1:** Demographic and deployment characteristics of the 81 confirmed CL cases.

| Characteristic | Sex | Military Service |  | Rank |  |  | Season deployed |  |
| --- | --- | --- | --- | --- | --- | --- | --- | --- |
|  | Male | Army | Navy | JNCO | SNCO | Ofr | Wet | Dry |
| **Number of individuals** | 81 | 77 | 4 | 69 | 7 | 5 | 58 | 23 |
| **%** | 100 | 95.1 | 4.9 | 85.2 | 8.6 | 6.2 | 71.6 | 28.4 |

Abbreviations: JNCO = junior non-commissioned officer; SNCO = senior non-commissioned officer; Ofr = officer

**Table 2:**
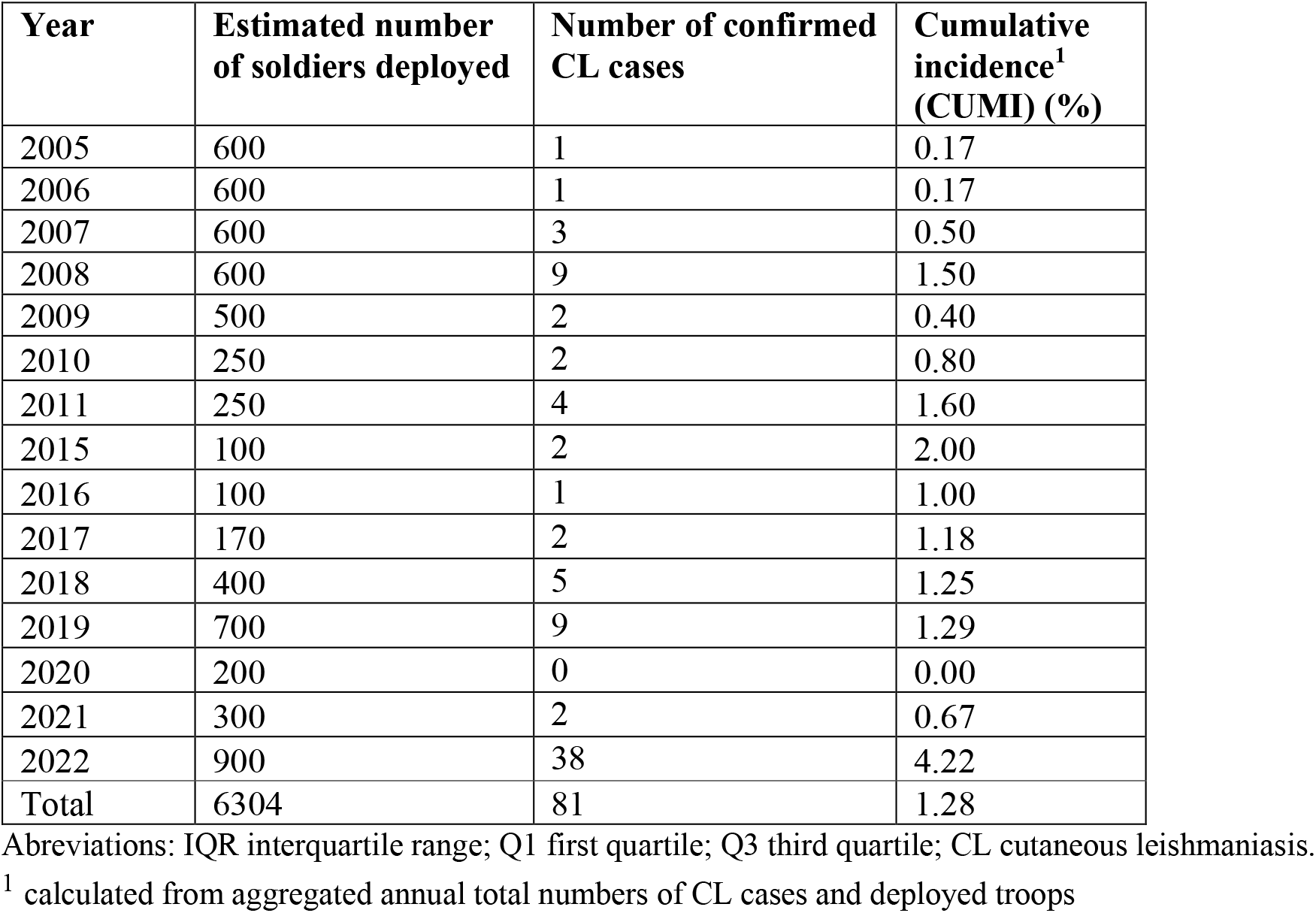
CL in British soldiers who participated in an 8–week JT in Belize from 2005 to 2022.

**Table 3:** Seasonal cumulative incidence (CUMI) of CL in British troop units deployed in an 8 – week jungle training in Belize between 2019 and 2022.

| Year | Number of units | Jungle training (JT) area | Month deployed | Season | Number of soldiers deployed | Number of confirmed CL cases | Cumulative incidence (CUMI) (%) |
| --- | --- | --- | --- | --- | --- | --- | --- |
| 2019 | 1<sup>st</sup> Unit | 1963 Lines | January | Dry | 100 | 0 | 0.0 |
|  | 2<sup>nd</sup> Unit | Chiquibul<sup>1</sup> | April | Dry | 120 | 0 | 0.0 |
|  | 3<sup>rd</sup> Unit | Gallon Jug<sup>1</sup> and Yalbac | September | Wet | 150 | 0 | 0.0 |
|  | 4<sup>th</sup> Unit | 1963 Lines | October | Wet | 330 | 9 | 2.7 |
| 2020 | 1<sup>st</sup> Unit | 1963 Lines | April | Dry | 80 | 0 | 0.0 |
|  | 2<sup>nd</sup> Unit | Yalbac | September | Wet | 120 | 0 | 0.0 |
| 2021 | 1<sup>st</sup> Unit | 1963 Lines | January | Dry | 120 | 0 | 0.0 |
|  | 2<sup>nd</sup> Unit | Gallon Jug<sup>1</sup> and Yalbac | September | Wet | 180 | 2 | 1.1 |
| 2022 | 1<sup>st</sup> Unit | Mountain Pine Ridge | January | Dry | 100 | 0 | 0.0 |
|  | 2<sup>nd</sup> Unit | 1963 Lines | April | Dry | 150 | 2 | 1.3 |
|  | 3<sup>rd</sup> Unit | Mountain Pine Ridge & Chiquibul<sup>1</sup> | September | Wet | 200 | 3 | 1.5 |
|  | 4<sup>th</sup> Unit | Gallon Jug<sup>1</sup> and Yalbac | October | Wet | 450 | 33 | 7.3 |

<sup>1</sup> JT areas where historical accounts report *Leishmania* infections in sand flies (see text for literature sources).

To investigate provision and adoption of PPMs to mitigate sand fly vector bite exposure during JT, a retrospective analysis was conducted on questionnaire responses collected from a cohort of 22 soldiers with confirmed CL during the recent outbreak in 2022. The questionnaire captured demographic information including sex, age, military rank, previous endemic overseas postings, and forest training area where billeted. Data described the breadth and depth of received PDHE regarding vector-borne diseases including leishmaniasis, utilisation of PPMs, and the soldiers’ attitudes and actual practices to mitigate exposure to biting insects.

Data were analysed using STATA v.18.

## 3. Results

### Cumulative incidence of CL

Between 2005 and 2022, a total of 6,304 British soldiers (predominantly males) participated in JT in Belize. Among these individuals, there were 81 confirmed cases of CL, all of whom were males. The median age of the cases was 25 years (range: 19–40 years). The majority of the cases were JNCOs (85.2%), in the army service (95.1%), and deployed to Belize during the wet season (71.6%) (Table 1). Due to the COVID-19 pandemic, only two units of limited numbers, were deployed in 2020 and 2021 (Table 3).

The median CUMI of CL between 2005 and 2021 was 0.9% (Q1–Q3: 0.34%–1.34%) with an annual variation of 0.2%-2.0% (Table 2; Fig 2). A positive upward trend in the CUMI with increasing year was detected, though this failed to reach statistical significance (Poisson regression: *b*=0.05, *z*=1.90, *P*=0.058). In 2022, the proportion of troops that acquired CL significantly increased from 43/5,370 pre-2022 to 38/900 (χ ^*2*^ =70.8, *P*<0.0001), with a CUMI of 4.2%. This represented a 5.3-fold increase in the relative risk of developing CL in 2022 (IRR=5.3 [95% C.I.s: 3.41, 8.16]), compared to the median CUMI for the years previous (*z*=7.47, *P*<0.0001).

**Figure 2:**
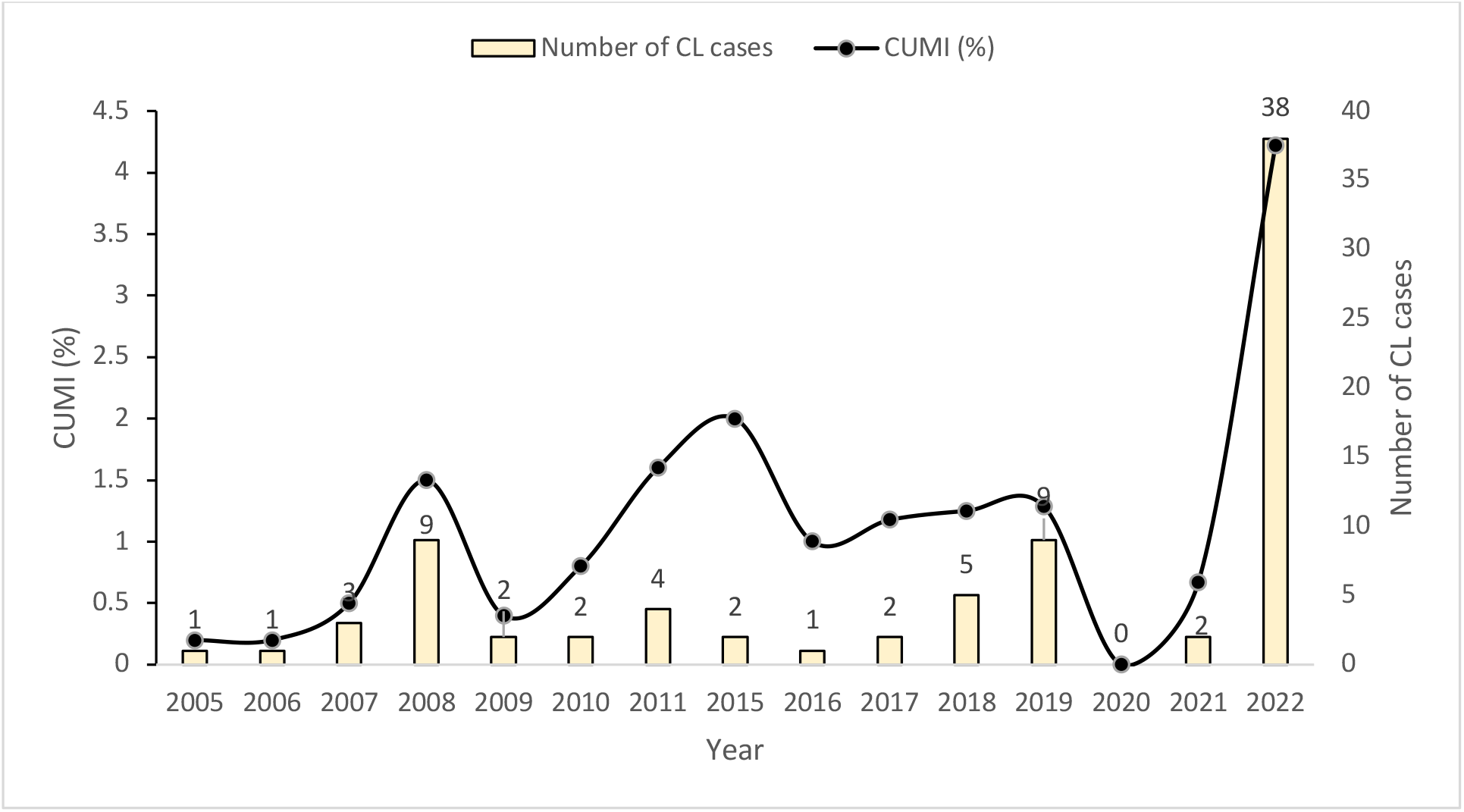
Cumulative incidence (CUMI) of CL in British troops following JT in Belize.

Higher resolution data were available for 2019 to 2022 (Table 3). Usually, four units on average undergo JT in Belize per year, two during the dry season and two during the wet season. The vast majority of CL cases (95.9%) during these years occurred among units that were deployed in the wet season. Most CL cases were associated with training in the 1963 Lines, Gallon Jug, and Yalbac JT areas, situated in the west of Belize (Fig 1).

Comparing seasons of troop deployment, the CUMI was significantly higher in the wet season (47/1,430) compared to in the dry season (2/670) (χ ^*2*^=17.88, *P*<0.001), even when excluding the unusually high number of CL cases in 2022 (11/780 vs 0/420) (χ ^*2*^=5.98, *P*=0.01) (Table 3).

### The outbreak of 2022

In late 2022, an outbreak involved 33 confirmed CL cases in a single unit of 450 men (CUMI=7.3%) (Table 3). This unit underwent JT in the wet season in Gallon Jug and Yalbac, otherwise known as the Orange Walk District (Fig 1), where past similar deployments, all in the wet season, resulted in 0/150 (2019) and 2/180 (2021) CL cases (Table 3). The 33 cases would have been exposed most likely during October 28^th^ to December 10^th^ during JT in 2021, and were initially documented from week 15 after their return to the UK, and continued to be confirmed up until week 26 (May 2022) (Fig 3). The median incubation period is uncertain as the precise dates of exposure to sand fly bites and subsequent infections were not observed. Taken from the last day of potential exposure, prior to departure from Belize (in week 6), to the time of lesion onset by self-reporting to the Medical Treatment Facility (MTF), the median incubation period would be 6.5 weeks (Q1–Q3: 4–12 weeks; range: 1–20 weeks). Alternatively, if it is assumed that exposure and transmission occurred during the mid-training period on average (week 3), the equivalent median incubation period could be 11 weeks (range: 1–22 weeks).

**Figure 3:**
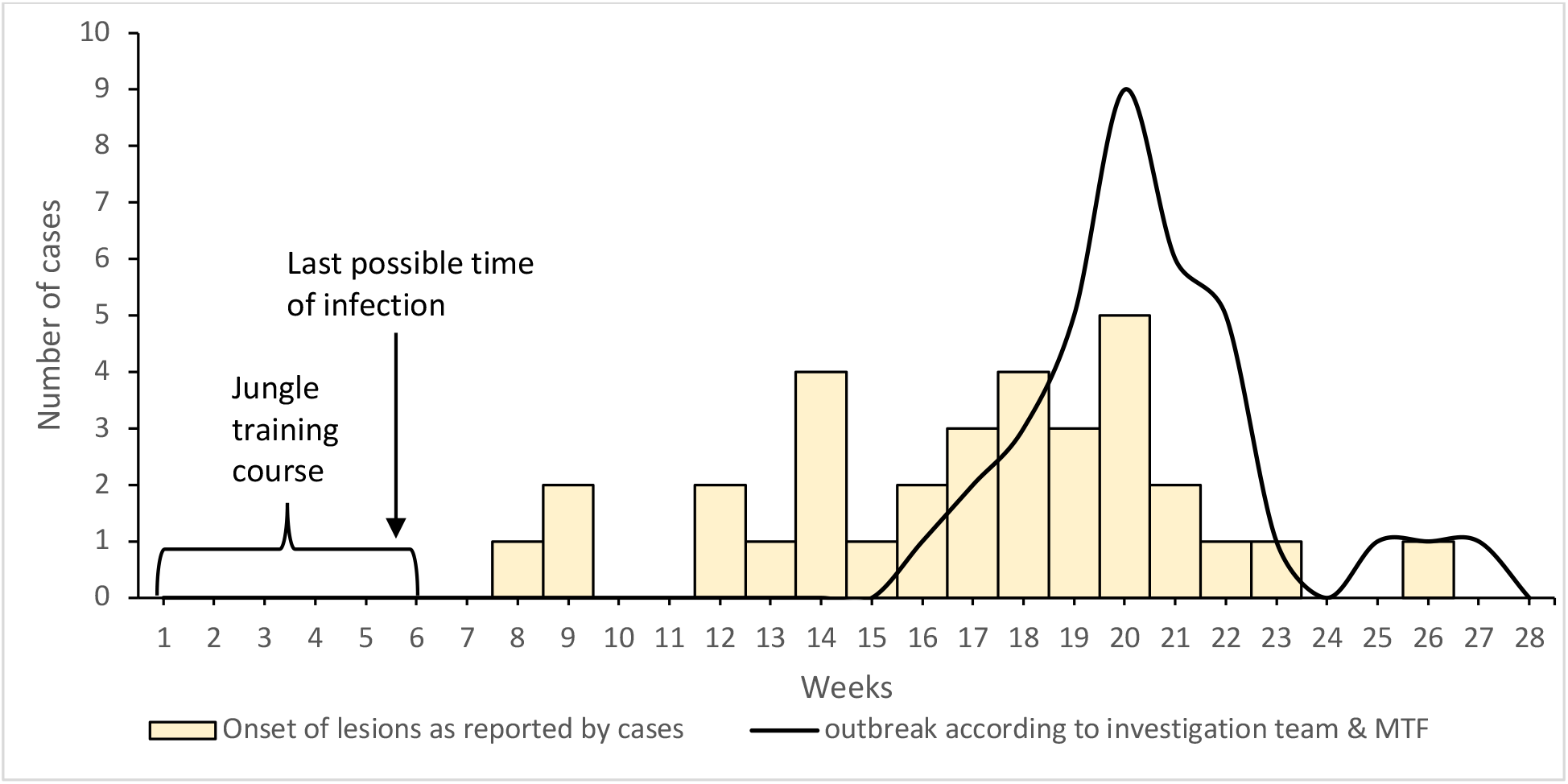
The numbers of CL cases that reported to the medical treatment facility (MTF) (weeks 16 to 27 post deployment) and the epidemic curve as described by the British military environmental health department.

### Mitigating practises

All 33 confirmed cases of the 2022 outbreak were sent a questionnaire. Of these, 22 individuals responded (a response rate of 66.7%). The demographic characteristics of the 22 cases were 100% male, and aged 18-41 years old (most [40.9%] 18-25 years old) (Table 4). The majority (68.2%) were JNCOs and had no previous overseas experience. Six of the soldiers had a history of overseas deployments to a *Leishmania*-endemic country, including Iraq, Belize, and Kenya in 2020 and 2021. None of the soldiers had a documented history of leishmaniasis prior to the 2022 outbreak.

**Table 4:**
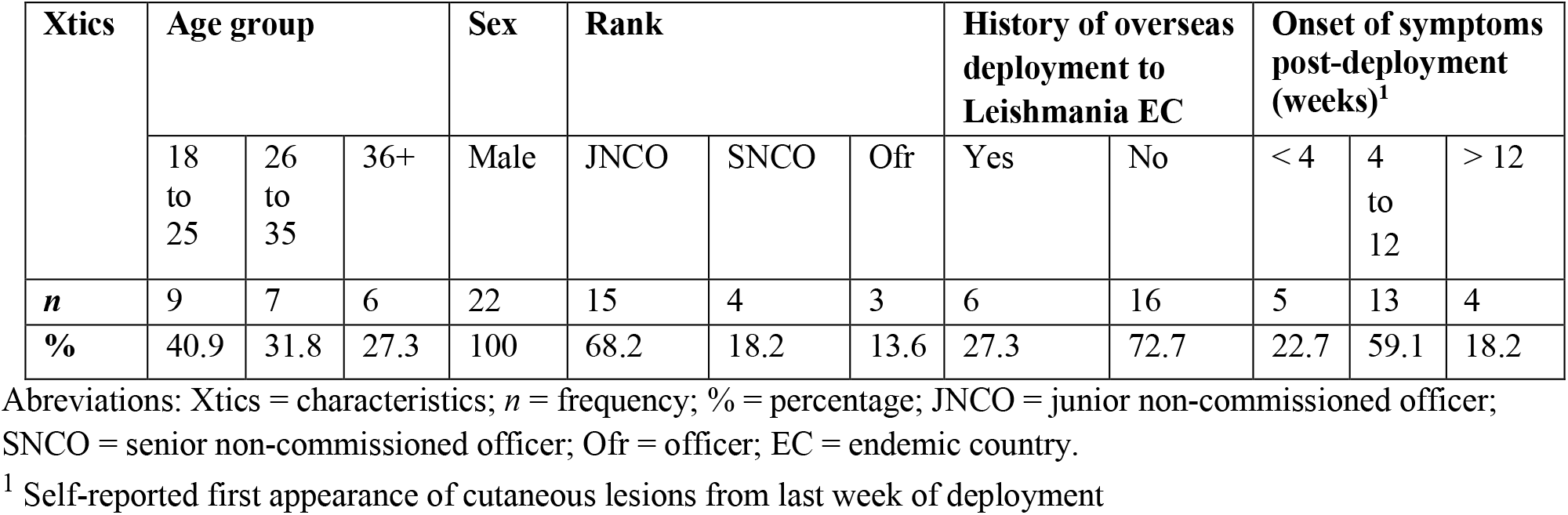
Demographic characteristics of 22 soldiers’ responses to the questionnaire.

### Pre-deployment health education (PDHE)

The majority 18/22 (81.8%) of respondants reported that they had received PDHE; one soldier did not, and three were unsure (Figure 4). Fourteen (63.6%) received PDHE from a CMT, and the remainder received from an EHP. These 18 soldiers reported receiving PDHE advice on PPMs including protective clothing and general methods to prevent insect bites. Only 6 (33.3%) of the 18 soldiers were specifically informed about sand flies and leishmaniasis, 4/6 delivered by an EHP (Figure 4).

**Figure 4:**
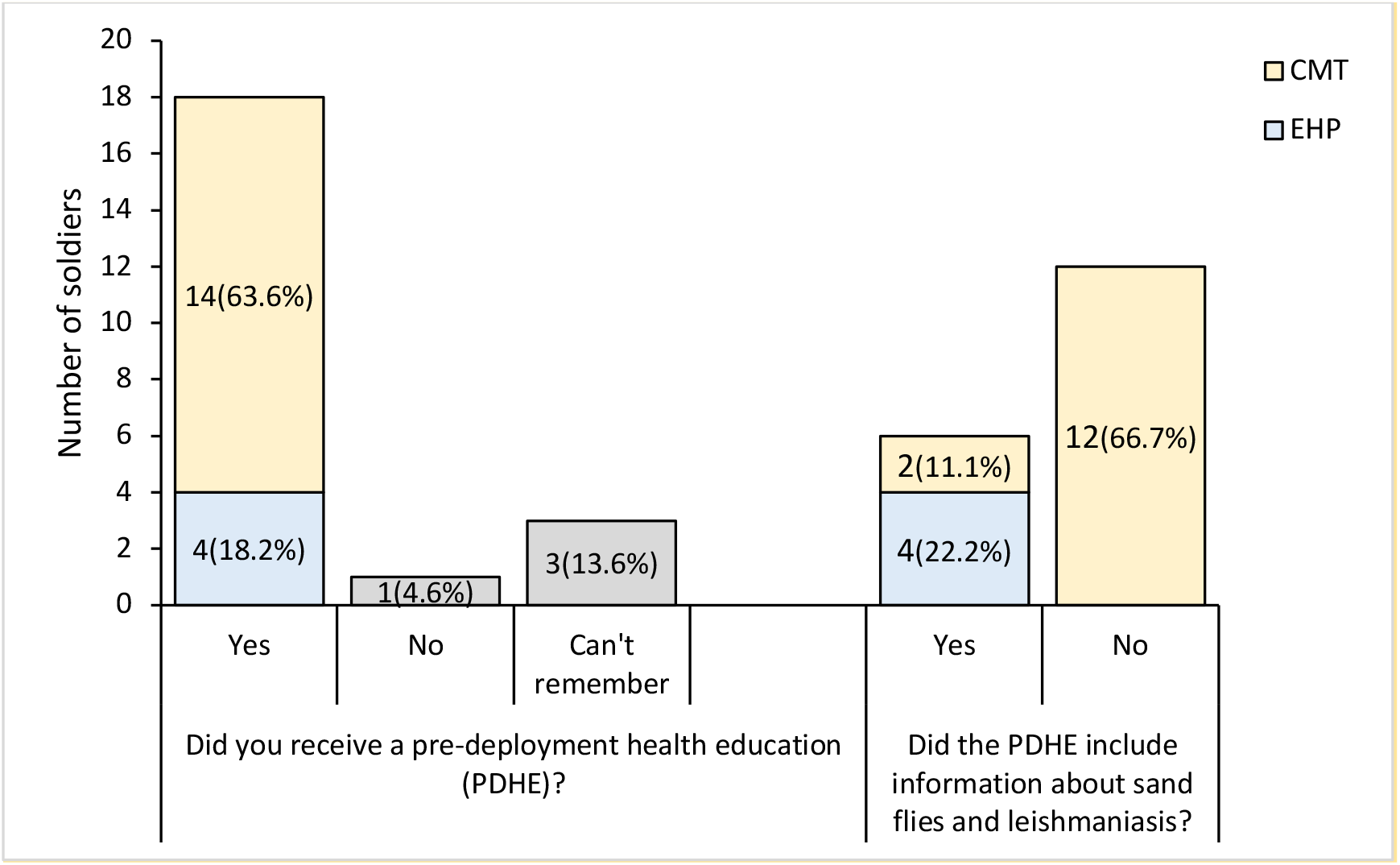
CL case responses to a questionnaire on pre-deployment health education (PDHE) including awareness of sand flies and leishmaniasis. The PDHE brief was delivered either by a CMT or an EHP.

### Provision and use of personal protective measures (PPMs) to avoid insect bites

All soldiers received hammocks before the jungle exercise (Ex) phase, but 10 (45.5%) reported the received hammocks were in poor condition (holes and/or tears) (Fig 5). Most (90.9%) of the soldiers treated their uniforms with Permapel (0.5% permethrin in 300ml) before the jungle Ex phase, whereas fewer (59.1%) treated their hammocks. Three (13.6%) soldiers requested new uniforms before their deployment (optional). All soldiers confirmed that they were provided with insect repellent (N,N-diethyl-meta-toluamide or DEET-34%) and head nets, however, over 40% and up to 90.9% did not use their insect repellent, wear their head nets, or wear long sleeved clothing consistently during the jungle Ex phase. Post jungle Ex phase, while on (R&R), 54.5% of the soldiers reported that they did not comply with these topical repellent or coverings (Figure 5).

**Figure 5:**
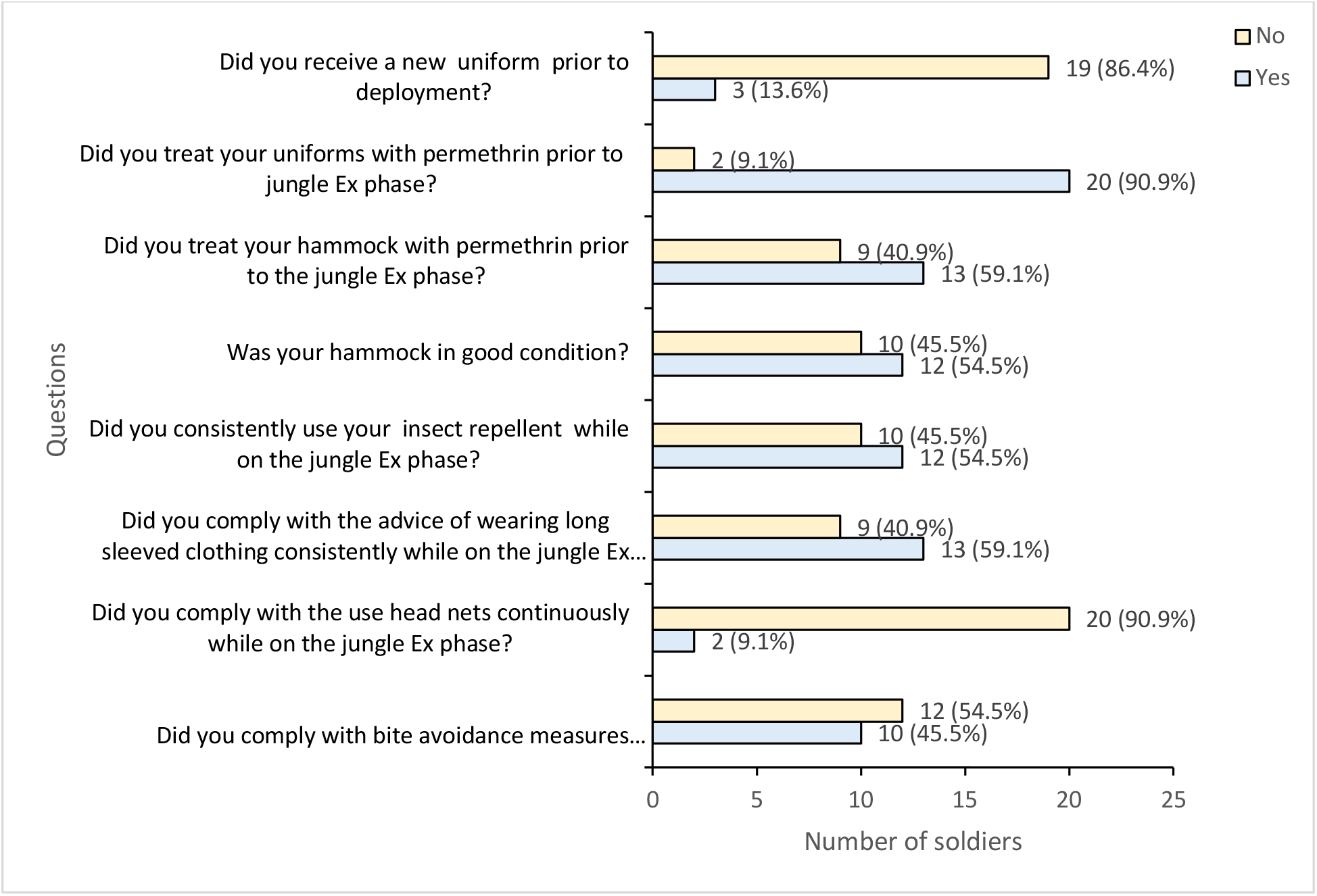
CL case responses to a questionnaire on the provision and compliance with personal protective measures (PPMs).

## 4. Discussion

The consequences of developing CL are not life-threatening, but at the same time are not trivial. Afflicted soldiers need to undergo lengthy treatments. This can be especially unpleasant particularly for infections with *L*. (*V*.) *braziliensis*, where prolonged/intensive treatments are necessary to clear the parasite in order to avoid serious disfigurement through the destruction of the naso-pharyngeal tissues, a condition known as mucocutaneous leishmaniasis, which often occurs only later in life. Second, the disease has financial implications to the armed forces. Chemotherapeutic treatments costs average £5,000 per patient, and uncomplicated cases are typically out of service for 8-24 weeks. Compensation claims against the MOD can be substantial. Thus, increasing vigilance, adoption of socially acceptable PPMs, and evaluation of complimentary vector control methods, are undoubtedly justified.

Whilst transmission risk occurs all year round, greater risk of infection (>95% of all CL cases and the CUMI) is experienced by troops deployed during the wet season compared to the dry season. The common sand fly species considered to be the important vectors in Belize include *Lutzomyia (Lu*.*) panamensis, Lu. shannoni, Lu. cruciatus*, and *Lu. ovallesi* [35]. All of these vector species are traditionally considered forest species where leaf-litter, tree buttresses and hollows and animal burrows are potential adult diurnal resting sites [34]. Information on the phenology of these sand fly species in Belize and elsewhere are scarce; many of the sand fly vectors in the Americas are more abundant during the wet season phases, whereas some are more dry-adapted, but spatio-temporal variations are reported [42].

Though not based on exhaustive investigations, historical accounts report *Leishmania* infections in *Lu. panamensis, Lu. shannoni, Lu. cruciatus, and Lu. ovallesi* sand flies in Gallon Jug, Chiquibul, and Guacamallo[34,35] where soldiers conduct JT today (Figure 1). The majority of CL cases among British soldiers between 2019 to 2022 occurred following JT in Gallon Jug, Yalbac, and 1963 Lines, all situated in the west of Belize. Belize lacks what is conventionally termed “primary” or “virgin” broadleaf forest, owing to historical land use practices by the ancient Maya peoples and various natural and human-induced disturbances like hurricanes, fires, and logging [43]. Consequently, areas untouched since 1980 are designated as “mature forests” rather than “primary forests” [44]. Although not as forested as the Toledo District in southern Belize [44], the western region of Belize is recognised for its mature forest, referred to locally as high bush [45]. This region habours the main JT areas (Gallon jug, Yalbac and Laguna Sector in the northwest, and Guacamallo in the southwest). Other JT areas such as Chiquibul and Mountain Pine Ridge have been described as medium bush, and Sibun Gorge as low bush [35]; all of which are mainly secondary forest. Hence it appears that sand fly vectors and *Leishmania* may be circulating in most if not all of these broadly characterised ecotypes (Table 3; Fig 1).

The primary aetiological agents of CL in the region are *L*. (*L*.) *mexicana* and *L*. (*V*.) *braziliensis*, the former being more prevalent [16–19,32,34]. The traditional occupational risk of CL due to *L. (L*.*) mexicana* infection (giving rise to the name chiclero’s ulcer) was predominant amongst collectors (“chicleros”) of chewing-gum latex (chiclé) from Sapodilla trees *Manilkara zapota*, conducted in forests during the wet season [46]. Beltran & Bustamente (1942) noted that most CL cases amongst 1,506 inhabitants of 58 surveyed chiclé camps in neighbouring Mexico occurred during the wet season, with CL prevalences of 16.7% and 2.2% in males and females, respectively [47]. An epidemiological survey of 14 geographically independent farming communities located in forested areas (n=1,448 people), revealed a similar 10.8% (18.1% males, 3.7% females) positive to *L. (L*.*) mexicana* skin test antigen which is indicative of exposure [48].

The 2022 military CL outbreak following training in western Belize represents a 5.3-fold (95% C.I.s: 3.41, 8.16) increase in the relative risk of developing CL compared to previous years. This increase is unexpectedly high, considering that the 1% CUMI is notably lower than the reported values in comparable military deployments in Belize (Dutch soldiers) [18], as well as the majority of values for French Guiana (French soldiers) [10] and Afghanistan (Dutch soldiers) [21], which share similar deployment characteristics (duration, training activities, and period).

The unsolved question is why did this outbreak occur? Plausible non-mutually exclusive reasons might include unusual climatic events promoting changes in sand fly vector and/or reservoir compositions or abundance affecting host-vector-parasite interactions; changes in troop susceptibility (e.g. host innate immunity or parasite virulence); and alterations in troop activities or behaviours which increase their exposure to infectious vector bites.

To initiate an investigation of the latter possiblity, the individuals afflicted during the 2022 outbreak in Unit 4 were asked for responses to a questionnaire covering the provision of, and compliance to, PPMs. The army provides PDHE through health briefings given to soldiers either prior to their deployment to Belize or immediately upon their arrival at BATSUB, and before the jungle Ex phase begins. The responses suggest that basic PDHE information was not effectively provided, and thus soldiers’ knowledge about sand flies and the risk of leishmaniasis was likely inadequate. PDHE sessions highlighting leishmaniasis risks and emphasising the importance of PPMs have been suggested as an effective way of preventing CL in military personnel in endemic regions [15,49,50]. In Colombia, knowledge about leishmaniasis was linked to an increased likelihood of soldiers adopting PPMs against insect bites [51]. Of course, knowledge about leishmaniasis alone does not guarantee the adoption and implementation of PPMs [51]. Failure to provide approriate protective equipment such as serviceable hammocks was reported. Notwithstanding, facilities were provided to reapply insect repellent (permethrin) to uniforms and hammocks prior to JT. Most of the soldiers (90.9%) did apply this treatment to their uniforms, but a substantial number (40.9%) failed to treat their hammocks. Studies indicate that reapplying topical permethrin to troop uniforms [52] and to hammocks [10] can effectively reduce the risk of sand fly contact and leishmaniasis.

Inconsistent use of long-sleeved clothing, head nets and DEET insect repellent during jungle Ex phase was also reported. Not regularly using the provided head nets was said to be due to discomfort, reduced visibility, heat, and facial irritation, which are in line with reasons provided by soldiers in other deployments [14]. Over 40% of soldiers did not consistently use provisioned DEET repellents and wear long-sleeved clothing during the jungle Ex phase, which is likely due to similar reasons. In one study, only 5% of soldiers regularly applied DEET repellent during training exercises [22]. The inconsistent use of DEET among military personnel is suggested to be a contributing factor to contracting leishmaniasis [14,18,20–24,52]. The social unacceptability of army formulated 34% DEET, as supplied to troops in Belize, is likely to be as much due to a lack of vector-borne disease awaremenss as due to its unpleasant presentation (repugnant smell, and accidential taste, and high viscosity). As commanders play a central role in establishing a culture of compliance and responsibility, command duty proves pivotal in promoting and enforcing the correct utilisation of protective measures among troops, as evidenced by a study of US troops deployed in various locations [22].

We acknowledge that the small sample in this study, and absence of controls, limits our ability to quantify and attribute relative risk to provision and adoption of PDHE and PPMs. Nonetheless, we consider the current data sufficient to initiate focussed investigations to improve delivery of PDHE and uptake of PPMs. Thus, the following recommendations are currently indicated, that (i) soldiers should receive adequate PDHE, preferably from professionals, emphasising transmission biology, the potential consequences of infection, and PPM options; (ii) soldiers should be equipped with all necessary and serviceable protective equipment prior to jungle Ex phase; and (iii) commanders should encourage and even enforce compliance. Further research is recommended to evaluate in more detail the effectiveness of PDHE on compliance and against vector exposure. More acceptable formulations of DEET-based insect repellent could be considered.

## Conclusions

CL, and the recent rise in case incidence, signifies a substantial burden to the British military. This preliminary study highlights the need for vigilance and preparedness e.g. mapping sand fly vector populations across training areas, to combat future CL outbreaks. Issues surround the provision and compliance to PPMs including the need for better communication and quality provisions are discussed.

## Data Availability

All data produced in the present work are contained in the manuscript

## Acknowledgements

We thank the Defence Public Health Unit and Capt Hale Isra for providing information on CL cases, and BATSUB for providing information on number of troops deployed over the study period. We also thank the outbreak investigation team (WO2 M. Soffe, SSgt MT Fatchu, Sgt S. Ward and Sgt M Ellis) who provided valuable information on the CL outbreak. This study was supported by the Research and Clinical Innovation pillar of the Defence Medical Services (RE: 22/23.003) and The Drummond Foundation (Horne/211122).

